# Homologous Ad26.COV2.S vaccination results in reduced boosting of humoral responses in hybrid immunity, but elicits antibodies of similar magnitude regardless of prior infection

**DOI:** 10.1101/2023.03.15.23287288

**Authors:** Thandeka Moyo-Gwete, Simone I. Richardson, Roanne Keeton, Tandile Hermanus, Holly Spencer, Nelia P. Manamela, Frances Ayres, Zanele Makhado, Thopisang Motlou, Marius B. Tincho, Ntombi Benede, Amkele Ngomti, Richard Baguma, Masego V. Chauke, Mathilda Mennen, Marguerite Adriaanse, Sango Skelem, Ameena Goga, Nigel Garrett, Linda-Gail Bekker, Glenda Gray, Ntobeko A.B. Ntusi, Catherine Riou, Wendy A. Burgers, Penny L. Moore

**Affiliations:** National Institute for Communicable Diseases of the National Health Laboratory Services, Johannesburg, South Africa; MRC Antibody Immunity Research Unit, School of Pathology, University of the Witwatersrand, Johannesburg, South Africa; Institute of Infectious Disease and Molecular Medicine, University of Cape Town, Observatory, South Africa; Division of Medical Virology, Department of Pathology; University of Cape Town; Observatory, South Africa; Department of Medicine, University of Cape Town and Groote Schuur Hospital; Observatory, South Africa; Cape Heart Institute, Faculty of Health Sciences, University of Cape Town; Observatory, South Africa; South African Medical Research Council Extramural Unit on Intersection of Non-communicable Diseases and Infectious Diseases, University of Cape Town, Cape Town, South Africa; South African Medical Research Council, Cape Town, South Africa; Centre for the AIDS Programme of Research in South Africa, Durban, South Africa; Discipline of Public Health Medicine, University of KwaZulu-Natal, Durban, South Africa; Desmond Tutu HIV Centre, Cape Town, South Africa; Wellcome Centre for Infectious Diseases Research in Africa, University of Cape Town, Observatory, South Africa

**Author notes:** these authors contributed equally to the work (Thandeka Moyo-Gwete, Simone I. Richardson, Roanne Keeton). Corresponding authors: Catherine Riou, Wendy Burgers and Penny L Moore.

**Keywords:** SARS-CoV-2, Ad26.COV2.S vaccine, antibodies, neutralization, ADCC, T cells, memory differentiation, hybrid immunity

## Abstract

The impact of previous SARS-CoV-2 infection on the durability of Ad26.COV2.S vaccine-elicited responses, and the effect of homologous boosting has not been well explored. We followed a cohort of healthcare workers for 6 months after receiving the Ad26.COV2.S vaccine and a further one month after they received an Ad26.COV2.S booster dose. We assessed longitudinal spike-specific antibody and T cell responses in individuals who had never had SARS-CoV-2 infection, compared to those who were infected with either the D614G or Beta variants prior to vaccination. Antibody and T cell responses elicited by the primary dose were durable against several variants of concern over the 6 month follow-up period, regardless of infection history. However, at 6 months after first vaccination, antibody binding, neutralization and ADCC were as much as 33-fold higher in individuals with hybrid immunity compared to those with no prior infection. Antibody cross-reactivity profiles of the previously infected groups were similar at 6 months, unlike at earlier time points suggesting that the effect of immune imprinting diminishes by 6 months. Importantly, an Ad26.COV2.S booster dose increased the magnitude of the antibody response in individuals with no prior infection to similar levels as those with previous infection.

The magnitude of spike T cell responses and proportion of T cell responders remained stable after homologous boosting, concomitant with a significant increase in long-lived early differentiated CD4 memory T cells. Thus, these data highlight that multiple antigen exposures, whether through infection and vaccination or vaccination alone, result in similar boosts after Ad26.COV2.S vaccination.

## Introduction

The Ad26.COV2.S vaccine, developed by Johnson and Johnson, is an adenovirus vector-based vaccine initially rolled out as a single dose regimen that conferred 85% efficacy against severe disease caused by SARS-CoV-2 (Sadoff et al., 2021a, 2021b). However, as variants of concern (VOCs) emerged, vaccine efficacy and effectiveness against infection and symptomatic disease decreased and hence, in South Africa, a booster dose was recommended (Feikin et al., 2022; Gray et al., 2022; Lin et al., 2022; Naranbhai et al., 2021; Rosenberg et al., 2021). A booster dose of the Ad26.COV2.S vaccine was reported to confer 75% protection against moderate to severe COVID-19 disease prior to the emergence of the Delta variant (Hardt et al., 2022) and this was consistent with the booster vaccine effectiveness of 72% against hospitalization during the Omicron BA.1 wave in South Africa (Gray et al., 2022).

As the pandemic progressed, the proportion of those with hybrid immunity increased substantially (Crotty, 2021). In South Africa, it is now estimated that the majority of individuals in the population have been infected with SARS-CoV-2 (Bingham et al., 2022; Madhi et al., 2022a; Sykes et al., 2021). We and others have shown that individuals infected with SARS-CoV-2 prior to, or following, vaccination have significantly increased humoral responses compared to those with infection or vaccination alone (Keeton et al., 2021; Kitchin et al., 2022; Reynolds et al., 2021; Stamatatos et al., 2021). Specifically, Ad26.COV2.S vaccinees who were previously infected with either D614G or Beta showed significantly boosted binding, neutralizing and ADCC responses. Conversely, prior SARS-CoV-2 infection did not significantly impact the magnitude of spike-specific-CD4 or CD8 T cell responses (Keeton et al., 2021). Similar results were obtained with ChAdOx nCov-19, another adenovirus-based vaccine, where a single dose of ChAdOx nCov-19 administered to participants previously infected with SARS-CoV-2 enhanced cross-reactive neutralizing antibody responses 1 month after vaccination (Chibwana et al., 2022; Madhi et al., 2022b).

Furthermore, “immune imprinting” has been reported, where the sequence of the initial antigen exposure determined the cross-reactivity of the antibody response regardless of subsequent heterologous exposures (Chemaitelly et al., 2022; Keeton et al., 2021; Rodda et al., 2022; Wheatley et al., 2021). In our previous study of Ad26.COV2.S vaccinated healthcare workers (HCWs), participants previously infected with the Beta variant had more cross-reactive antibody responses compared to those previously infected with the D614G variant (Keeton et al., 2021).

Although a number of studies have now assessed the durability of immune responses to the Ad26.CoV.2S vaccine, confirming that Ad26.CoV.2S-induced antibody and T cell responses remain stable over an 8-month period, the impact of previous infection with different variants on the durability of the immune response has not been well described (Barouch et al., 2021; Collier et al., 2021; Khoo et al., 2022; Tan et al., 2022; Zhang et al., 2022). In this study, we longitudinally assessed antibody and T cell responses elicited by Ad26.COV2.S vaccination in a previously described cohort of HCWs who were either SARS-CoV-2 naïve, or had been infected with the D614G or Beta variants prior to vaccination (Keeton et al., 2021). We first defined the magnitude of spike-specific immune responses up to 6 months after vaccination and found these varied in titer, but were durable after a single Ad26.COV2.S dose, regardless of prior infection status. Secondly, we show that a homologous Ad26.COV2.S boost increased the magnitude of the antibody response to similar levels regardless of prior infection status, or the infecting variant, and expanded the proportion of early differentiated memory CD4 T cells specific for spike. Determining the durability, magnitude and boosting potential of immune responses in the context of hybrid immunity, now near universal, remains key to help inform policy on the type and frequency of booster administration, and to better understand immune protection against SARS-CoV-2.

## Results

### Antibody and T cell responses elicited by Ad26.COV2.S vaccination are maintained for up to 6 months regardless of prior infection

To determine the durability of immune responses following a single dose of Ad26.COV2.S vaccination, we examined longitudinal antibody and T cell responses in a previously-described cohort of HCWs (Keeton *et al*., 2021). The cohort included COVID-19 naïve participants (Group 1) and participants who had either a D614G (Group 2) or Beta (Group 3) infection prior to Ad26.COV2.S vaccination (**Fig. S1A**). The median time between SARS-CoV-2 infection and vaccination was 7.2 months for Group 2 (interquartile range (IQR): 6.6-8.6) and 2.4 months for Group 3 (IQR: 1.8-2.7). Blood was drawn 22 days (IQR: 14-29) prior to vaccination for all groups (pre-vax samples) and approximately 1 month (median of 29 days, IQR: 28-34) after vaccination. A subset of these participants (n=13, 14 and 16 for groups 1, 2 and 3, respectively) had follow-up samples taken at approximately 6 months after vaccination enabling us to perform durability studies (**Fig. S1A; Fig. S1B**). Of these, 6, 10 and 8 participants (groups 1, 2 and 3, respectively) received a Ad26.COV2.S booster dose approximately 9 months after the first vaccination (median of 8.8, IQR: 8.7-8.9) and blood was drawn at a median of 23 days (IQR: 21-24) after the vaccine boost (**Fig. S1A; Fig. S1B**). As breakthrough infection (BTI) dramatically impacts antibody kinetics and T cell function (Bates et al., 2022; Kitchin et al., 2022), any participants with detectable BTIs were excluded from these analyses (n=9).

We first longitudinally assessed the binding antibody response to D614G using a SARS-CoV-2 spike enzyme linked immunosorbent assay (ELISA), antibody dependent cellular cytotoxicity (ADCC) responses using a FcγRIIIa activation assay and neutralizing antibodies using a pseudovirus-based assay (**Fig. 1A-C**). Antibody binding responses against D614G significantly increased 1 month after vaccination and remained high at month 6 in all groups. As expected, geometric mean (GM) values were higher in the previously infected groups compared to Group 1 at all tested time points (**Fig. 1A**).

**Figure 1.**
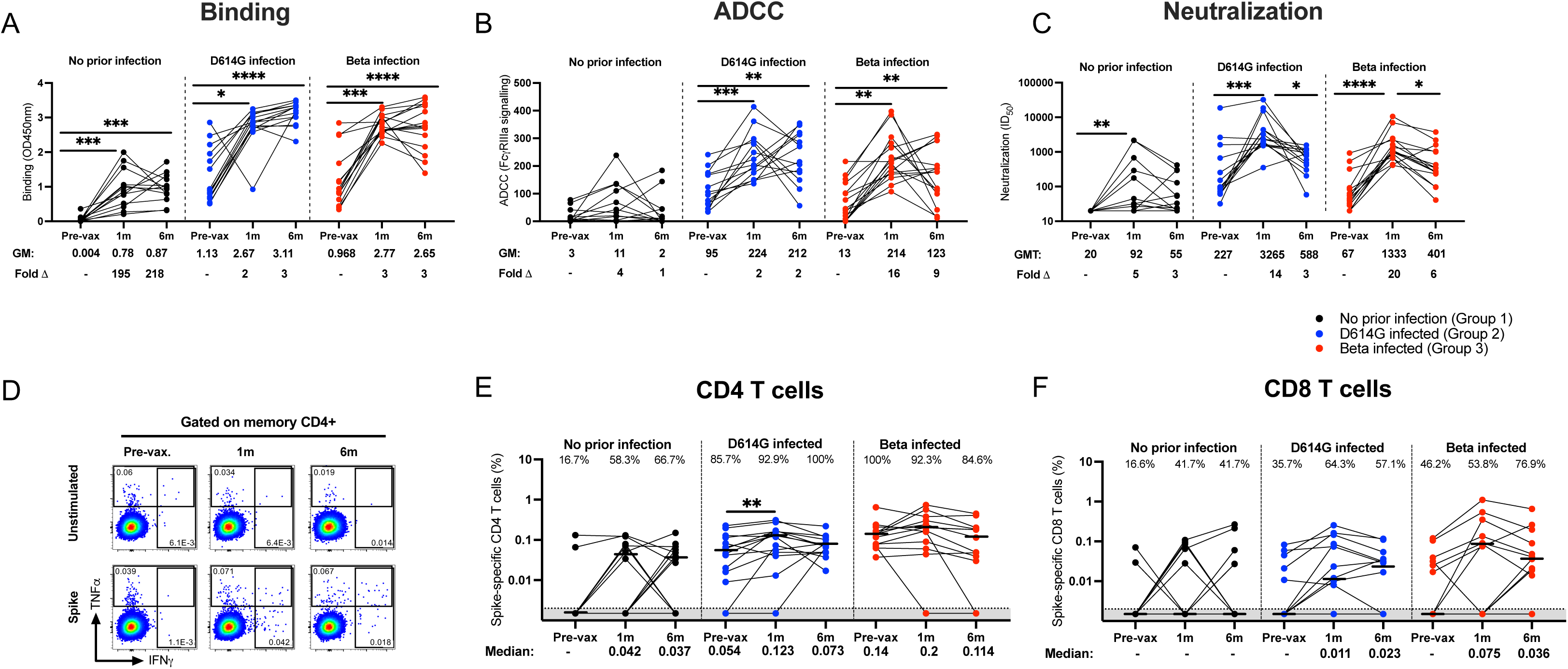
Durability of antibody and T cell responses 6 months after a single Ad26.COV2.S dose. Participants with no prior infection (black, Group 1), infected with D614G (blue, Group 2) or infected with Beta (red, Group 3) were tested for binding (**A**), antibody-dependent cellular cytotoxicity (ADCC) activity (**B**), and neutralization (**C**) of the D614G variant. Ancestral spike-specific CD4 (**D and E**) and CD8 (**F**) T cell responses before vaccination and at 1 and 6 months after vaccination. For antibody responses, geometric mean (GM) values and fold changes are shown below the graphs. Fold changes were all calculated relative to the pre-vaccination timepoints. Binding antibodies were quantified by OD450nm values, ADCC activity was measured in FcγRIIIa signaling and neutralization was indicated by 50% inhibitory dilution (ID_50_) values. All serology experiments were performed in duplicate. For T cell responses, the proportion of responders (%) is shown within the graph. Frequencies are reported after background subtraction and medians are indicated below the graph. Representative flow plots are shown in **D.** The Friedman test with Dunn’s correction for multiple comparisons was used to determine statistical significance. Significance is shown as: ****p<0.0001, ***<0.001, **p<0.01, *p<0.05.

For ADCC, only participants with prior infection showed a statistically significant increase at 1 month post-vaccination. Infection-naïve participants in Group 1 showed a 4-fold increase from baseline, but this was statistically insignificant (**Fig. 1B**). While ADCC was maintained between 1 and 6 months in Groups 2 and 3 overall, there was considerable variation within the groups, with a substantial proportion of individuals either showing increased or decreased ADCC over this period. This heterogeneity was more pronounced than for the other functions we measured, likely implicating other modulators of Fc effector function including antibody isotype or subclass and glycosylation (Lu et al., 2018).

Neutralizing antibody responses against D614G were significantly increased in all three groups at 1 month after vaccination. Although reduced D614G neutralizing antibody responses were observed between 1 and 6 months post-vaccination (Group 1: geometric mean titer (GMT) of 92 vs 55; Group 2: 3,265 vs 588 and Group 3: 1,333 vs 401, respectively), neutralizing activity remained detectable in most of the samples tested (**Fig. 1C).** Overall, all three antibody functions were relatively durable against D614G with a slight drop in neutralizing titers and ADCC by 6 months post-vaccination. Antibody levels were significantly higher in the previously infected groups compared to SARS-CoV-2-uninfected participants at each time point tested (**Fig.1A-C**).

Next, we assessed the durability of spike-specific T cell responses over a 6 month period by measuring IFN-γ, IL-2 and TNF-α cytokine production after stimulation with a peptide pool targeting ancestral spike (**Fig. 1D and S2A**). As shown previously (Keeton et al., 2021), all groups mounted robust spike-specific CD4 and CD8 T cell responses after primary vaccination (**Fig. 1E** and **F**). Spike-specific CD4 T cell response frequencies were well maintained at 6 months post-vaccination in all groups (median: 0.042 vs 0.037, 0.123 vs 0.073, and 0.2 vs 0.114 for groups 1, 2 and 3 respectively) (**Fig. 1E**). No appreciable change was detected in the proportion of CD4 T cell responders between 1 and 6 months after the initial vaccination in any of the tested groups.

The magnitude of the SARS-CoV-2-specific CD8 T cell response detected at 1 month after vaccination was maintained at 6 months for all groups with no significant reduction in the proportion of responders for Groups 1 and 2 at 6 months (41.7% and 64.3% vs 41.7% and 57.1%, respectively). Interestingly, a 23% increase in responders was detected at 6 months in Group 3 (53.8% at 1 month vs 76.9% at 6 months; **Fig. 1F**). We also noted that five participants across the 3 groups gained and four lost their CD4 response and a further five gained and four lost a CD8 response at the 6 month time point (**Fig. 1E** and **F**). Overall, these results demonstrate that the robust CD4 and CD8 T cell responses generated after primary vaccination are detectable 6 months later.

### Hybrid immunity confers high levels of cross-reactivity 6 months after vaccination, regardless of the infecting variant

We next compared antibody functions against SARS-CoV-2 variants D614G, Beta, Delta and Omicron (BA.1) across the three groups 6 months after vaccination to assess the degree and durability of cross-reactivity (**Fig. 2A-C; Fig. S3; Fig. S4; Fig. S5**). The degree of cross-reactivity for each group is summarized in **Fig 2D**, which shows the GMT against D614G, Beta, Delta and Omicron variants and SARS-CoV-1 for binding antibodies, neutralization and ADCC. Although the overall dynamics of antibody responses mirrored those observed for D614G, titers against the VOCs and SARS-CoV-1 were much lower as expected **(Fig. S3; Fig. S4; Fig. S5)**. Binding responses were significantly higher in the previously infected Groups 2 and 3, compared to Group 1, with up to 5-fold differences in binding potency (**Fig. 2A**). This is likely an underestimate of these fold changes, as binding antibody values in Groups 2 and 3 reached the upper limit of detection of the assay. The previously infected groups also displayed substantially elevated ADCC and neutralization activity towards SARS-CoV-2 variants compared to Group 1 (up to 33-fold and 19-fold, respectively) (**Fig. 2B; Fig. 2C**). Interestingly, despite differences in the infecting variants and the time between infection and vaccination (7 and 2 months, respectively) before vaccination, Group 2 and Group 3 had similar levels of all three antibody functions (**Fig. 2A-C; Fig. 2D**) except for titers against SARS-CoV-1, which trended higher for Group 3 compared to Group 2 (**Fig. 2C; Fig. 2D**). Thus, while early immune imprinting resulted in slightly different patterns of cross-reactivity at 1 month post-vaccination (Keeton et al., 2021), these differences did not persist at 6 months after vaccination in this cohort.

**Figure 2:**
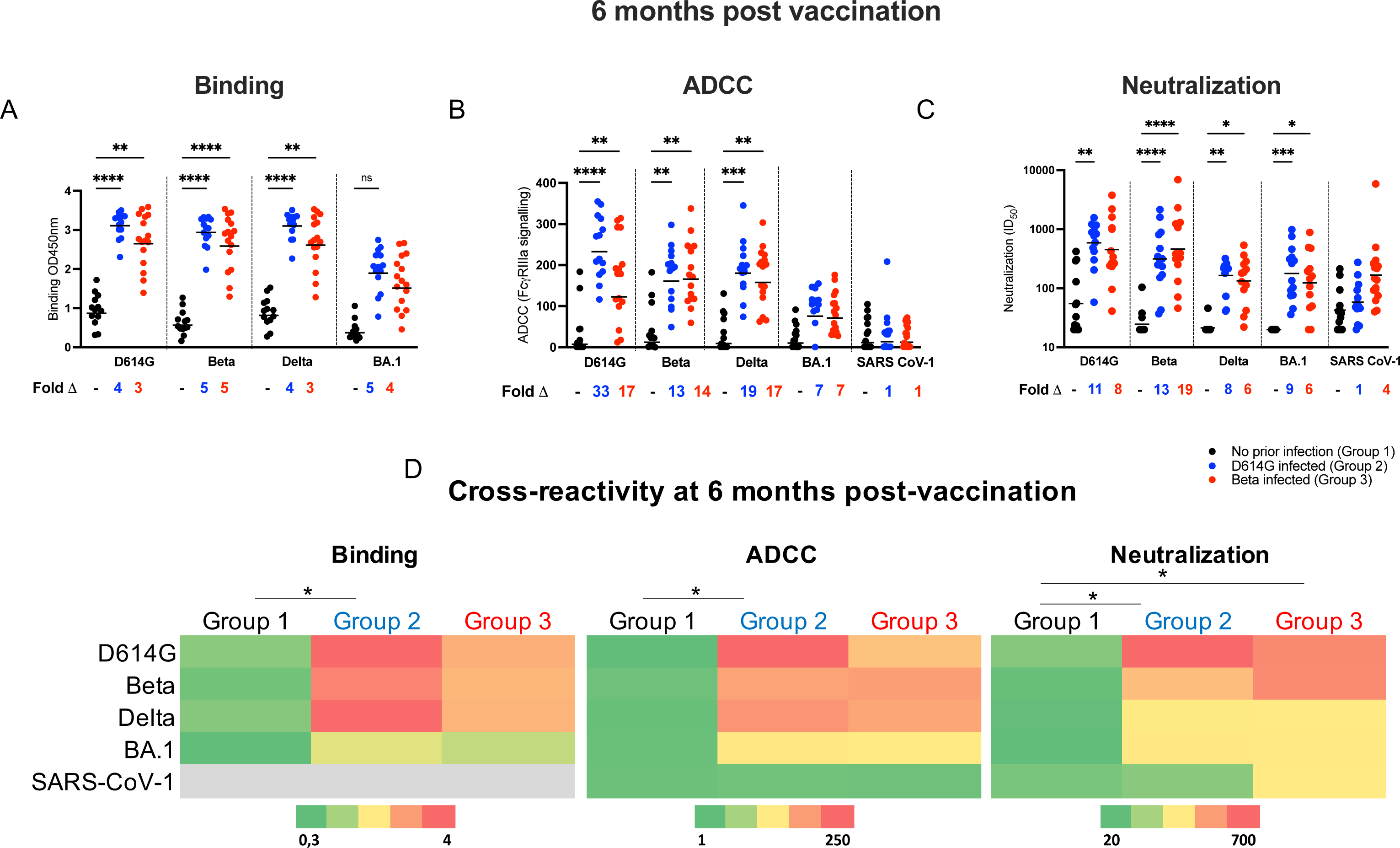
Cross-reactivity of vaccinee plasma samples at 6 months post vaccination. Plasma was tested for cross-reactivity in antibody binding (**A**), ADCC activity (**B**) and neutralization assays (**C**) against D614G, Beta, Delta, Omicron (BA.1) and SARS-CoV-1 for participants with no prior infection (black), participants infected with D614G (blue) or Beta infected participants (red). Fold changes between participant groups are shown below the graphs. Binding antibodies were quantified by OD450nm values, ADCC activity was measured in FcγRIIIa signaling and neutralization was indicated by ID_50_ values. (**D**) Heat-map depicting cross-reactivity of the plasma. The scale depicted shows strongest responses in red and weakest responses in green for each assay. All experiments were performed in duplicate. Statistical significance between the different groups was determined using the Kruskal-Wallis test with Dunn’s correction for multiple comparisons. Significance is shown as: ****p<0.0001, ***<0.001, **p<0.01, *p<0.05.

### Prior infection dampens the boosting of antibody responses

Approximately 9 months after the initial Ad26.COV2.S dose, a subset of participants received an additional Ad26.COV2.S vaccine (n=6, 10 and 8 in Group 1, 2 and 3, respectively, **Fig. S1A**). Antibody responses against the D614G variant measured 6 months after the first dose were compared to those measured one month after the boost (**Fig. 3**). In the SARS-CoV-2 naïve Group 1 participants, boosting with the Ad26.COV2.S vaccine resulted in a 2-fold increase in binding activity (**Fig. 3A**), 14-fold increase in ADCC activity (**Fig. 3B**) and 16-fold increase in neutralization titers (**Fig. 3C**). The fold increase was lower in participants with prior infection, with no change in binding antibodies, a 3-fold increase in ADCC in groups 2 and 3 and a 6-7 fold increase in neutralizing activity in participants with prior infection after a Ad26.COV2.S booster (**Fig. 3B; Fig. 3C**). Importantly, one month after the vaccine boost, the antibody binding titer and ADCC activity levels were comparable across all groups, irrespective of the SARS-CoV-2 variant tested (**Fig. 3D-E)**. Although not statistically significant, neutralization potency against the Beta and BA.1 variants was higher in Groups 2 and 3 compared to Group 1 (6- and 10-fold for Beta and 4- to 6-fold for BA.1, respectively) (**Fig. 3F**).

**Figure 3.**
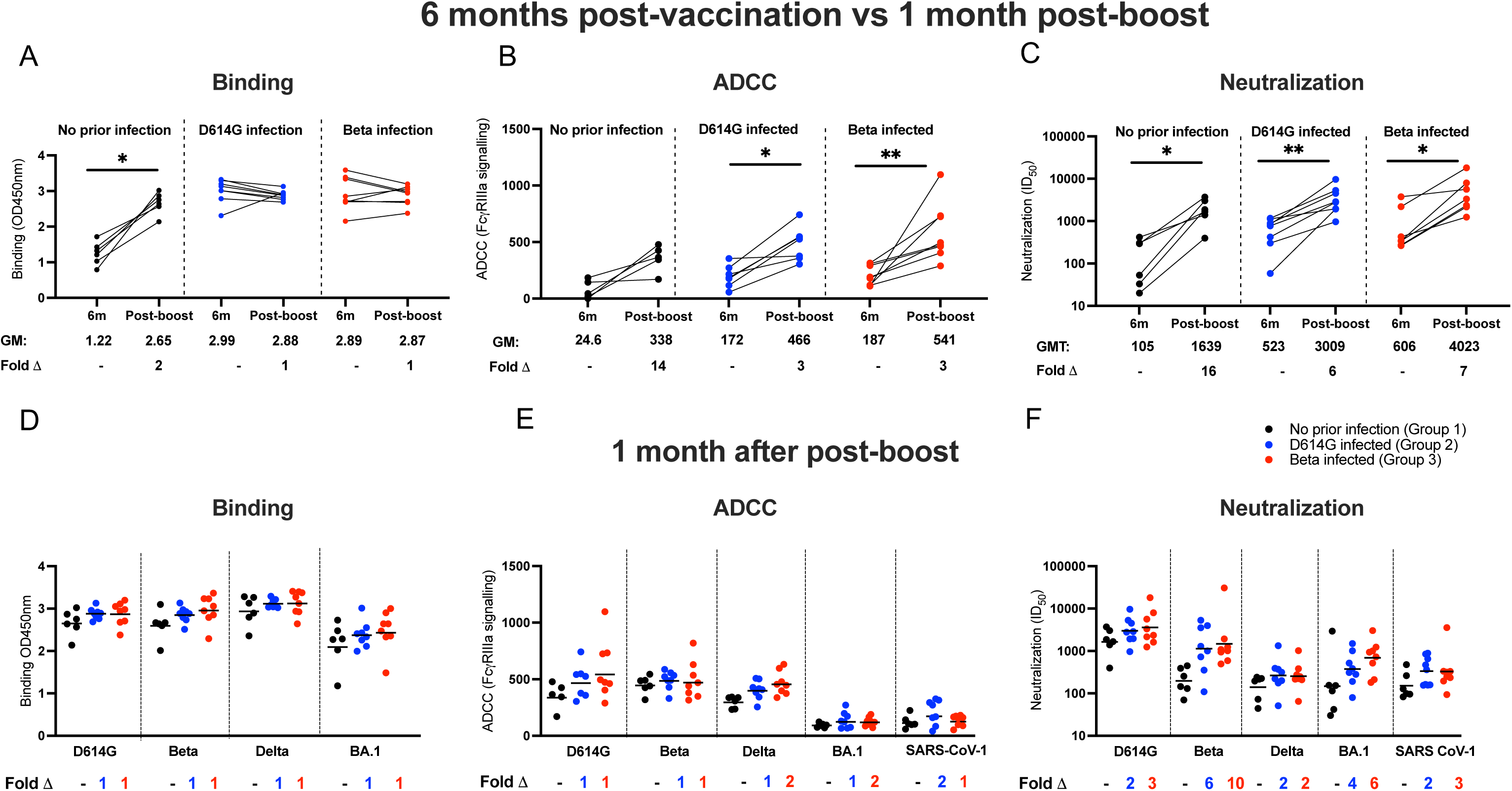
The antibody response after an Ad26.COV2.S booster dose. Antibody responses were measured at 6 months post-vaccination and compared to responses at one month after a homologous boost. Plasma samples from patients with either no prior infection (black), infected with D614G (blue), and infected with Beta (red) were tested for binding (**A**), antibody-dependent cellular cytotoxicity (ADCC) activity (**B**) and neutralization (**C**) to the D614G variant. Geometric mean values are shown below the graphs. Binding antibodies were quantified by OD450nm values, ADCC activity was measured in FcγRIIIa signaling and neutralization was indicated by ID_50_ values. All experiments were performed in duplicate. The Wilcoxon matched-pairs signed rank test was used to determine statistical significance between the 6 months post-vaccination and one month post-boost time points. Significance is shown as: **p<0.01, *p<0.05. Binding (**D**), ADCC activity (**E**) and neutralizing (**F**) responses between groups were tested one month after the booster dose. Plasma activity was tested against the D614G, Beta, Delta and BA.1 variants and SARS-CoV-1. Fold changes are shown below the graphs. All experiments were performed in duplicate. Statistical significance between the different groups was determined using the Kruskal-Wallis test with Dunn’s correction for multiple comparisons. None of the comparisons were statistically significant.

### Ad26.COV2.S boost expands spike-specific early differentiated memory T cells

To determine the effect of a homologous vaccine booster on T cell immunity, we compared the magnitude and memory differentiation phenotype of CD4 and CD8 T cells 6 months after the primary Ad26.COV2.S vaccine and 1 month after the boost in participants for which PBMC samples were available at both time points (n= 5, 10 and 5 for groups 1, 2, and 3, respectively) (**Fig. 4A-C**). No significant changes were detected in the magnitude of the spike-specific T cell response or the proportion of responders for either CD4 or CD8 T cells before and after boosting in any of the groups. There was however some heterogeneity at the individual level after boosting, with 4/20 participants across all groups having CD4 T cell responses demonstrating increased magnitudes and two gaining a response. Similarly, CD8 T cells had higher cytokine responses in 4/20 participants and three gained a response (**Fig. 4B; Fig. 4C**).

**Figure 4.**
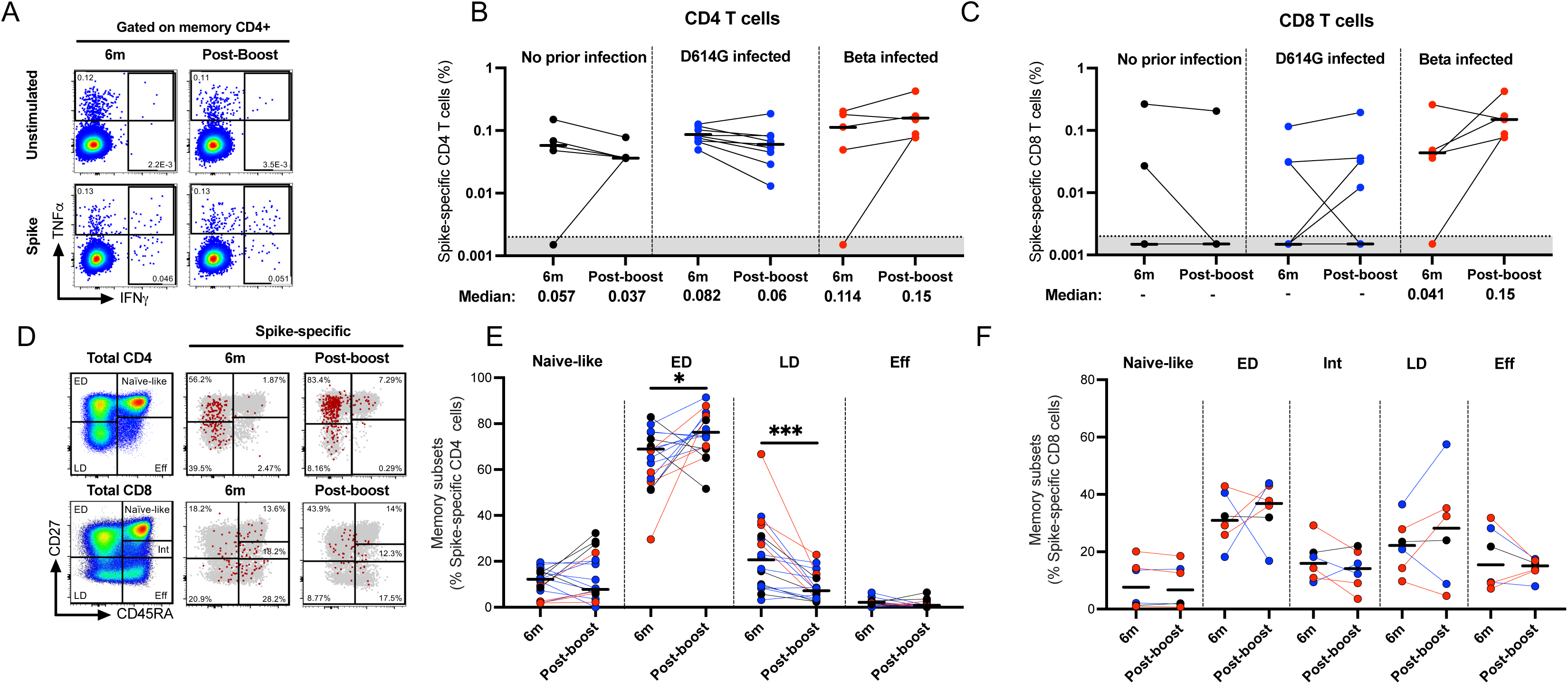
T cell response and phenotyping after an Ad26.COV2.S booster dose. T cell cytokine responses were measured at 6 months post-vaccination and compared to responses at one month after a homologous boost. PBMCs from patients with either no prior infection (black), infected with D614G (blue), or infected with Beta (red) were tested for frequency of spike-specific CD4+ (**A and B**) and CD8+ T cells (**C**) producing any of the studied cytokines (IFN-γ, IL-2 and TNF-α) in response to ancestral peptide pools. Bars represent medians with values indicated below the graphs. Data are plotted as background subtracted total cytokine responses. Due to high TNF-α background, single TNF-α producing cells were excluded from analysis. Representative plots are shown in **A**. Representative overlay flow plots of CD27 and CD45RA expression are shown in **D**. Red dots depict SARS-CoV-2-spike-specific expression at 6 months (middle) and post-boost (right). Grey dots represent total CD4 (upper panel) or CD8 T cells (lower panel). Using CD27 and CD45RA, four memory subsets can be delineated (left panel): naive (CD27+CD45RA+), early differentiated (ED; CD27+CD45RA-), late differentiated (LD; CD27-CD45RA-) and effector (Eff; CD27-CD45RA+) subsets. Summary graphs of the frequency (%) of each subset within SARS-CoV-2-spike-specific CD4 (**E**) or CD8 T cells (**F**). Wilcoxon matched-pairs signed rank test was used to determine statistical significance. Significance is shown as: ***<0.001, **p<0.01, *p<0.05.

To investigate the phenotypic characteristics of spike-specific T cell responses, we analyzed their memory differentiation profiles (**Fig. 4D-F; Fig. S2B-D**). The measurement of CD45RA and CD27 enabled the detection of four distinct CD4 T cell populations, namely naïve-like (CD45RA+CD27+), early differentiated (ED: CD45RA-CD27+), late differentiated (LD: CD45RA-CD27-) and effector (Eff: CD45RA+CD27-) populations, with CD8 T cells having a fifth population of intermediate cells (Int: CD45RA+CD27low) (**Fig. 4D**). Due to the low number of responders meeting the threshold for phenotypic analysis, we did a bulk analysis combining the three groups. The memory profile of the spike-specific CD4 T cells after the booster vaccination displayed a significant increase in ED (p = 0.032) and a concomitant decrease in LD (p < 0.0001) subsets compared to the 6-month time-point (median: 68.2% vs 74.9% and 21.9% vs 7.2% respectively; **Fig. 4E**). No significant differences were detected among the CD8 T cell memory populations (**Fig. 4F**).

## Discussion

Although a single dose of the Ad26.COV2.S vaccine has been shown to induce durable adaptive immune responses (Barouch et al., 2021; Tan et al., 2022), the effect of previous infection on the titer and longevity of these responses and the effect of a homologous boost have not been fully explored. Our data show that while antibody activity elicited by a single Ad26.COV2.S dose was durable up to 6 months after initial vaccination, Fc functionality, antibody neutralization and binding titers remained significantly lower in individuals with no prior infection compared to individuals with hybrid immunity. A homologous vaccine booster resulted in all antibody responses reaching a similar magnitude irrespective of prior SARS-CoV-2 infection, however the magnitude of boosting in the context of hybrid immunity was reduced compared to that of previously uninfected participants. Both CD4 and CD8 SARS-CoV-2 spike-specific T cell responses were sustained 6 months after vaccination in all groups. However, unlike the antibody responses, a homologous boost did not increase the magnitude or proportion of T cell responders but did expand an early differentiated CD4 memory subset. These data suggest that boosting with the Ad26.COV2.S vaccine is beneficial for increasing antibody titers in individuals with no history of prior infection and inducing long-lived CD4 memory T cells in all groups.

We and others have shown that breakthrough infection after vaccination yields substantially higher neutralization, binding and Fc effector functions compared to vaccination alone (Bates et al., 2022; Kitchin et al., 2022; Walls et al., 2021) and leads to better protection from hospitalization and severe disease (Bobrovitz et al., 2023). Here, we extend our previous observation that infection prior to Ad26.COV2.S vaccination results in superior responses compared to vaccination alone. This latter form of hybrid immunity is likely the most relevant current immune scenario for South Africa, which has extremely high population seropositivity (Bingham et al., 2022; Madhi et al., 2022a; Sykes et al., 2021).

Interestingly, our study differs from the ChAdOx1 nCoV-19 vaccine where individuals who were seropositive at the start of the study did not have increased neutralizing titers after the second dose of the vaccine (Gelanew et al., 2022; Madhi et al., 2022b). The ChAdOx1 nCoV-19 vaccine, also an adenovirus-based vaccine but not based on the stabilized spike protein, elicited antibodies that were maintained for up to 6 months post-vaccination in seropositive individuals, but large drops were observed in seronegative individuals (Madhi et al., 2022b). In our study, both baseline seronegative and seropositive groups maintained their antibody levels. This may be a consequence of the stabilized spike in combination with the adenoviral delivery system used in the Ad26.COV2.S vaccine inducing a more durable response, differences in infection timing or participant characteristics between the studies.

We found that in individuals with prior infection, ADCC is only slightly boosted after a single Ad26.COV2.S dose. Additionally, ADCC shows more heterogeneity in decay of activity compared to neutralization over 6 months in vaccinees with prior infection. Others have found that the Fc response is not only quantitatively higher for individuals who received mRNA vaccination with prior infection but also qualitatively superior (Bowman et al., 2022). While known to occur following Ad26.COV2.S administration, Fc effector function has not previously been assessed longitudinally or in the context of Ad26.COV2.S hybrid immunity (Alter et al., 2021; Richardson et al., 2022). This study focused on binding, neutralization and ADCC, but there are several additional antibody functions that could be examined in the context of hybrid immunity, including antibody dependent cellular phagocytosis and complement deposition. Therefore, assessing whether our findings are applicable beyond ADCC for adenoviral vectors would be beneficial.

Different SARS-CoV-2 variants have been shown to elicit varying immune responses, which may also impact the quantity and quality of the subsequent vaccine-induced responses (Dupont et al., 2021; Keeton et al., 2021; Moyo-Gwete et al., 2021; van der Straten et al., 2022). This phenomenon, known as “immune imprinting”, has been widely reported for influenza infection and vaccination (Belongia et al., 2016; Fonville et al., 2014). Previously, we reported that prior infection one month after Ad26.COV2.S vaccine primes the immune system in different ways, depending on the infecting variant (Keeton et al., 2021). In this study, we saw slightly higher neutralization of Beta and BA.1 in Group 2 (infected with Beta) one month after a homologous boost. However, overall, we did not find strong evidence for immune imprinting leading to differing cross-reactivity at 6 months after vaccination in individuals who were infected with the D614G or Beta variants prior to vaccination. This may be a consequence of antibody maturation by somatic hypermutation leading to broadening of responses regardless of the prior infecting variant. There is evidence of continual somatic hypermutation as a result of persistent viral antigen in the gut after SARS-CoV-2 infection as well as persistence of germinal centers for up to 3 months after BNT162b2 vaccine (Gaebler et al., 2021; Turner et al., 2021; Wang et al., 2020).

Analysis of the durability of the SARS-CoV-2 specific T cell response highlights that hybrid immunity consisting of D614G or Beta infection in combination with Ad26.COV2.S vaccination does not confer an advantage with respect to T cell responses over a 6 month period. This is supported by a similar study which demonstrated equivalent magnitude and durability of T cell responses after mRNA vaccination in individuals with or without prior infection (Goel et al., 2021).

Ad26.COV2.S boosting also did not significantly increase the T cell response in the three groups tested, in agreement with the findings of others (Atmar et al., 2022; Khoo et al., 2022). This is in keeping with our recent findings demonstrating that increasing numbers of SARS-CoV-2 exposures, whether through vaccinations or infections, do not result in sequential increases in the magnitude of the spike-specific T cell response (Keeton et al., 2022). Emerging data on the SARS-CoV-2 spike-specific T cell immune responses to different booster vaccination strategies suggest that heterologous boosting may offer an advantage over homologous boosting, with numerous studies now showing an increased T cell response when an mRNA boost is administered after the Ad26.COV2.S primary vaccine (Atmar et al., 2022; Khoo et al., 2022; Sablerolles et al., 2022).

Despite the lack of enhanced T cell cytokine response frequencies stimulated by Ad26.COV2.S booster vaccination, we did observe an expansion of early differentiated memory T cells, a subset likely to confer a long-lived response.

Another study demonstrated that after a primary mRNA vaccination regimen, early differentiated effector memory 1 (EM1) T cells (CCR7+,CD27+,CD45RA-) were maintained over a 6 month period (Goel et al., 2021). Notably, they found that a high proportion of EM1 T cells in the peak CD4 T cell response was associated with increased durability of the overall T cell response. This highlights the contribution of early differentiated EM1 T cells to the long-lived memory CD4 T cell response and supports the notion that the increased early differentiated memory T cell population (CD27+, CD45RA-) observed in our study after booster vaccination may result in an increase in the durability of the CD4 T cell response in these boosted participants.

Altogether, in the context of countries such as South Africa where, either through infection or vaccination, SARS-CoV-2 seropositivity is extremely high in some areas (Bingham et al., 2022), a booster vaccination for individuals who received one dose of Ad26.COV2.S regardless of prior infection status is likely to still be beneficial to the humoral and cellular response.

## Methods

### Cohort Description

Healthcare workers (HCWs; n=400) were enrolled in a longitudinal study from Groote Schuur Hospital (Cape Town, Western Cape, South Africa) as described previously (Keeton et al., 2021). Briefly, forty-three participants were included in this study who fell into one of three groups: (i) No evidence of previous SARS-CoV-2 infection by diagnostic PCR test or serial serology; (ii) infection during the ‘first wave’ of the pandemic in South Africa, prior to 1 September 2020, with known date of PCR-confirmed SARS-CoV-2 infection; and (iii) infection during the ‘second wave’, with known date of PCR-confirmed SARS-CoV-2 infection between 1 November 2020 and 31 January 2021.

### SARS-CoV-2 spike and nucleocapsid enzyme linked immunosorbent assay

SARS-CoV-2 D614G, Beta, Delta and Omicron (BA.1) variant spike proteins were expressed in Human Embryonic Kidney (HEK) 293F suspension cells by transfecting the cells with the spike plasmid. After incubating for six days at 37°C, 70% humidity and 10% CO_2_, proteins were first purified using a nickel resin followed by size-exclusion chromatography. Relevant fractions were collected and frozen at -80°C until use. Two µg/mL of spike or nucleocapsid (BioTech Africa; Catalogue number: BA25-P) protein were used to coat 96-well, high-binding plates and incubated overnight at 4°C. The plates were incubated in a blocking buffer made up of 1x PBS, 5% skimmed milk powder, 0.05% Tween 20. Plasma samples were diluted to 1:100 in a blocking buffer and added to the plates. Secondary antibody was diluted to 1:3000 in blocking buffer and added to the plates followed by TMB substrate (Thermo Fisher Scientific). Upon stopping the reaction with 1 M H_2_SO_4_, absorbance was measured at a 450nm wavelength. Antibodies CR3022, BD23, P2B-2F6 and 084-7D were used as positive controls and to differentiate between variants. The human anti-nucleocapsid 1A6 antibody (Thermo Fisher Scientific; Catalogue number: MA535941) was used as a positive control for the nucleocapsid ELISA. Palivizumab was used as a negative control in all experiments.

### Pseudovirus neutralization assay

SARS-CoV-2 pseudotyped lentiviruses were prepared by co-transfecting the HEK293T cell line with either SARS-CoV-2 D614G, Beta (L18F, D80A, D215G, K417N, E484K, N501Y, D614G, A701V, Δ242-244), Delta (T19R, R158G L452R, T478K, D614G, P681R, D950N, 156-157 del) or BA.1 (Omicron) (A67V, Δ69-70, T95I, G142D, Δ143-145, Δ211, L212I, 214EPE, G339D, S371L, S373P, S375F, K417N, N440K, G446S, S477N, T478K, E484A, Q493R, G496S, Q498R, N501Y, Y505H, T547K, D614G, H655Y, N679K, P681H, N764K, D796Y, N856K, Q954H, N969K, L981F) spike plasmids in conjunction with a firefly luciferase encoding lentivirus backbone plasmid. For the neutralization assay, heat-inactivated plasma samples from vaccine recipients were incubated with the SARS-CoV-2 pseudotyped virus for 1 h at 37°C, 5% CO_2_. Subsequently, 1x10^4^ HEK293T cells engineered to overexpress ACE-2 were added and incubated at 37°C, 5% CO_2_ for 72 h upon which the luminescence of the luciferase gene was measured. CB6, CA1 and 084-7D were used as positive controls and to differentiate between variants.

### FcγRIIIa activation (Antibody dependent cellular cytotoxicity) assay

The ability of plasma antibodies to cross-link *FcγRIIIa* and spike expressing cells was measured as a proxy for antibody dependent cellular cytotoxicity (ADCC). HEK293T cells were transfected with 5 µg of SARS-CoV-2 original variant spike (D614G), Beta, Delta or Omicron (BA.1) spike plasmids and incubated for 2 days at 37°C. Expression of spike was confirmed by binding of antibodies CR3022, P2B-2F6 and AIRU946-A6 and their detection by anti-IgG APC staining measured by flow cytometry. Subsequently, 1x10^5^ spike transfected cells per well were incubated with heat inactivated plasma (1:100 final dilution) or control mAbs (final concentration of 100 µg/mL) in RPMI 1640 media supplemented with 10% FBS 1% Pen/Strep (GIBCO, Gaithersburg, MD) for 1 h at 37°C. Jurkat-Lucia NFAT-CD16 cells were added and incubated for 24 h at 37°C, 5% CO_2_. Twenty µL of supernatant was then transferred to a white 96-well plate with 50 *μ*L of reconstituted QUANTI-Luc secreted luciferase and read immediately. Relative light units (RLU) of a no antibody control were subtracted as background. Palivizumab was used as a negative control, CR3022 and AIRU946-A6 as positive controls and P2B-2Fb to differentiate variants. To induce the transgene, 1x cell stimulation cocktail (Thermo Fisher Scientific) and 2 µg/mL ionomycin in R10 was added as a positive control. RLUs for D614G, Beta, Delta and BA.1 variants were normalized using CR3022 and AIRU946-A6, which bind similarly to the variants.

### Isolation of PBMC

Blood from heparin tubes was processed within 3 h of collection. Peripheral blood mononuclear cells (PBMC) were isolated using Ficoll-Paque by density gradient sedimentation (Amersham Biosciences, Little Chalfont, UK) as per the manufacturer’s instructions and cryopreserved in freezing media consisting of heat-inactivated fetal bovine serum (FBS, Thermo Fisher Scientific) containing 10% DMSO and stored in liquid nitrogen until use.

### Cell stimulation and flow cytometry staining

Cryopreserved PBMC were thawed, washed and rested in RPMI 1640 containing 10% heat-inactivated FCS for 4 h, seeded in a 96-well V-bottom plate at ∼2 × 106 PBMC per well and stimulated with SARS-CoV-2 spike peptide pools at a final concentration of 1 μg/mL (PepTivator®, Miltenyi Biotech, Bergisch Gladbach, Germany). These peptides were a combination of two commercially available peptide pools (PepTivator®, Miltenyi Biotech, Bergisch Gladbach, Germany) and are 15-mer sequences with 11 amino acids (aa) overlap based on the Wuhan-1 strain. They cover the N-terminal S1 domain of SARS-CoV-2 from aa 1 to 692, as well as the majority of the C-terminal S2 domain. All stimulations were performed in the presence of Brefeldin A (10 μg/mL, Sigma-Aldrich, St Louis, MO, USA) and co-stimulatory antibodies against CD28 (clone 28.2) and CD49d (clone L25) (1 μg/mL each; BD Biosciences, San Jose, CA, USA). As a negative control, PBMC were incubated with co-stimulatory antibodies, Brefeldin A and an equimolar amount of DMSO.

Cells were stimulated for 16 h, washed and stained with LIVE/DEAD Fixable VIVID Stain (Invitrogen, Carlsbad, CA, USA) prior to surface staining with the following antibodies: CD14 Pac Blue (TuK4, Invitrogen Thermo Fisher Scientific), CD19 Pac Blue (SJ25-C1, Invitrogen Thermo Fisher Scientific), CD4 PerCP-Cy5.5 (L200, BD Biosciences, San Jose, CA, USA), CD8 BV510 (RPA-8, Biolegend, San Diego, CA, USA), CD27 PE-Cy5 (1A4, Beckman Coulter), CD45RA BV570 (HI100, Biolegend, San Diego, CA, USA). Cells were then fixed and permeabilized using a Cytofix/Cyto perm buffer (BD Biosciences) and stained with CD3 BV650 (OKT3) IFN-γ Alexa 700 (B27), TNF-α BV786 (Mab11) and IL-2 APC (MQ1-17H12) from Biolegend. Finally, cells were washed and fixed in CellFIX (BD Biosciences). Samples were acquired on a BD LSR-II flow cytometer and analyzed using FlowJo (v10, FlowJo LLC, Ashland, OR, USA). A median of 272 380 CD4 events (IQR: 216 796 - 355 414) and 147 695 CD8 events (IQR: 109 697 - 202 204) were acquired. For cytokine analysis, cells were gated on singlets, CD14- CD19-, live lymphocytes, CD4 and CD8 and memory cells (excluding naive CD27+ CD45RA+ population). Results are expressed as the frequency of CD4+ or CD8+ memory T cells expressing IFN-γ, TNF-α or IL-2. For memory phenotyping, cells were gated on singlets, CD14- CD19-, live lymphocytes, CD4 and CD8 followed by IFN-γ, TNF-α and IL-2 with memory cells being gated on the IFN-γ or IL-2 subset. To ensure the robustness of phenotyping, we applied a cut- off of 40 SARS-CoV-2 specific T cell cytokine events. Due to high TNF-α background levels, cells producing TNF-α alone were excluded from the analysis.

## Declarations

### Ethics approval and consent to participate

The study was approved by the University of Cape Town Human Research Ethics Committee (Ethics number: HREC 190/2020 and 209/2020) and the University of the Witwatersrand Human Research Medical Ethics Committee (Ethics number: M210429). Written informed consent was obtained from all participants.

## Consent for publication

All authors have provided consent for publication.

## Availability of data and materials

All data are readily available upon request to the corresponding authors.

## Competing interests

All authors declare no competing interests.

## Funding

Research reported in this publication was supported by the South African Medical Research Council (SA MRC) with funds received from the South African Department of Science and Innovation (DSI), including grants 96825, SHIPNCD 76756 and DST/CON 0250/2012. This work was also supported by the Poliomyelitis Research Foundation (21/65) and the Wellcome Centre for Infectious Diseases Research in Africa (CIDRI-Africa), which is supported by core funding from Wellcome Trust (203135/Z/16/Z and 222574). We acknowledge funding from the Bill and Melinda Gates Foundation, through the Global Immunology and Immune Sequencing for Epidemic Response (GIISER) program. PLM is supported by the South African Research Chairs Initiative of the Department of Science and Innovation and National Research Foundation of South Africa (NRF 9834), the SA Medical Research Council SHIP program, the Centre for the AIDS Programme of Research in South Africa (CAPRISA). TMG is funded by a South African Medical Research Council Self-Initiated Research Grant. SIR is funded by the Poliomyelitis Research Foundation.

WAB and CR are supported by the EDCTP2 programme of the European Union’s Horizon 2020 programme (TMA2017SF-1951-TB-SPEC to C.R. and TMA2016SF- 1535-CaTCH-22 to WAB) and the Wellcome Trust (226137/Z/22/ Z). CR is supported by the National Institutes of Health (NIH) (R21AI148027). NABN acknowledges funding from the SA MRC, MRC UK, NRF and the Lily and Ernst Hausmann Trust.

## Author Contributions

TMG, SIR, RK, CR, WAB and PLM designed the study and wrote the manuscript. TMG, SIR, RK, TH, CR and NPM analyzed the data. TH, NPM, FA, ZM, HS and TM performed antibody assays. RK, MBT, NB, AN, RB and MVC performed T cell assays. MM, MA and SS managed the HCW cohort and contributed clinical samples. AG, NG, L-GB, and GG established and led the Sisonke vaccine study. NABN and WAB established and led the HCW cohort. All authors reviewed and approved the manuscript prior to submission.

## Data Availability

All data are readily available upon request to the corresponding authors.

## Acknowledgments

We acknowledge the participants who volunteered for this study. The SARS-CoV-2 Wuhan spike was provided by Jason McLellan (University of Texas) and parental pseudovirus plasmids by Elise Landais and Devin Sok (IAVI). We thank Bronwen E. Lambson for making the pseudovirus constructs.

## Supplementary figure legends

**Supplementary Figure 1. Study design and participant demographics.** (**A and B**) Schematic representing the study design and participant demographics. Participants were categorized into three groups: Group 1: no infection prior to vaccination (n=13), Group 2: D614G-infected prior to vaccination (n=14) and Group 3: Beta-infected prior to vaccination (n=16). All three groups were followed for 6 months after initial vaccination. A subset of n=6 Group 1, n=10 Group 2 and n=8 Group 3 participants received a homologous Ad26.COV2.S vaccination and boosting effect was analyzed after 1 month. Schematic was created using BioRender.com.

**Supplementary Figure 2. Flow cytometry gating strategy.** Gating strategy **(A and B)** and representative examples of SARS-CoV-2 spike-specific IFN-γ, IL-2 and TNF- α production in memory CD4+ and CD8+ T cells **(C)**. Representative plots showing spike-specific T cell immune phenotyping **(D)**. ED: early differentiated, LD: late differentiated, Eff: effector, Inter: intermediate.

**Supplementary Figure 3. Durability of binding responses 6 months after a single dose of Ad26.COV2.S.** Plasma samples from participants with no prior infection (black), infected with D614G (blue) or infected with Beta (red) were tested for binding responses to the Beta (**A**), Delta (**B**) and BA.1 (**C**) variants at three time points (pre-vaccination, 1 and 6 months post-vaccination). Geometric mean values and median fold changes are shown below the graphs. Fold changes were all calculated relative to the pre-vaccination time points. Binding antibodies were quantified by OD450nm values. All experiments were performed in duplicate. The Friedman test with Dunn’s correction for multiple comparisons was used to determine statistical significance. Significance is shown as: ****p<0.0001, ***<0.001, **p<0.01, *p<0.05.

**Supplementary Figure 4. Durability of antibody dependent cellular cytotoxicity 6 months after a single dose of Ad26.COV2.S.** Plasma samples from participants with no prior infection (black), infected with D614G (blue) or infected with Beta (red) were tested for ADCC activity to the Beta (**A**), Delta (**B**), BA.1 (**C**) variants and SARS-CoV-1 (**D**) at three time points (i.e, pre-vaccination, 1 and 6 months post-vaccination). Geometric mean values and median fold changes are shown below the graphs. Fold changes were all calculated relative to the pre-vaccination timepoints. ADCC activity was measured in FcγRIIIa signaling. All experiments were performed in duplicate. The Friedman test with Dunn’s correction for multiple comparisons was used to determine statistical significance. Significance is shown as: ****p<0.0001, ***<0.001, **p<0.01, *p<0.05.

**Supplementary Figure 5. Durability of neutralizing responses 6 months after single Ad26.COV2.S dose.** Plasma samples from participants with either no prior infection (black), infected with D614G (blue), and infected with Beta (red) were tested for neutralizing responses to the Beta (**A**), Delta (**B**), BA.1 (**C**) variants and SARS-CoV-1 (**D**). Plasma was tested over three time points; pre-vaccination, 1 and 6 months post-vaccination. Geometric mean values and fold changes are shown below the graphs. Fold changes were all calculated relative to the pre-vaccination timepoints. Neutralization was indicated by ID_50_ values. All experiments were performed in duplicate. The Friedman test with Dunn’s correction for multiple comparisons was used to determine statistical significance. Significance is shown as: ****p<0.0001, ***<0.001, **p<0.01, *p<0.05.

## Notes

### Competing Interest Statement

The authors have declared no competing interest.

