## Supplemental Data Figures 1-5 for "Homologous Ad26.COV2.S vaccination results in reduced boosting of humoral responses in hybrid immunity, but elicits antibodies of similar magnitude regardless of prior infection"

A

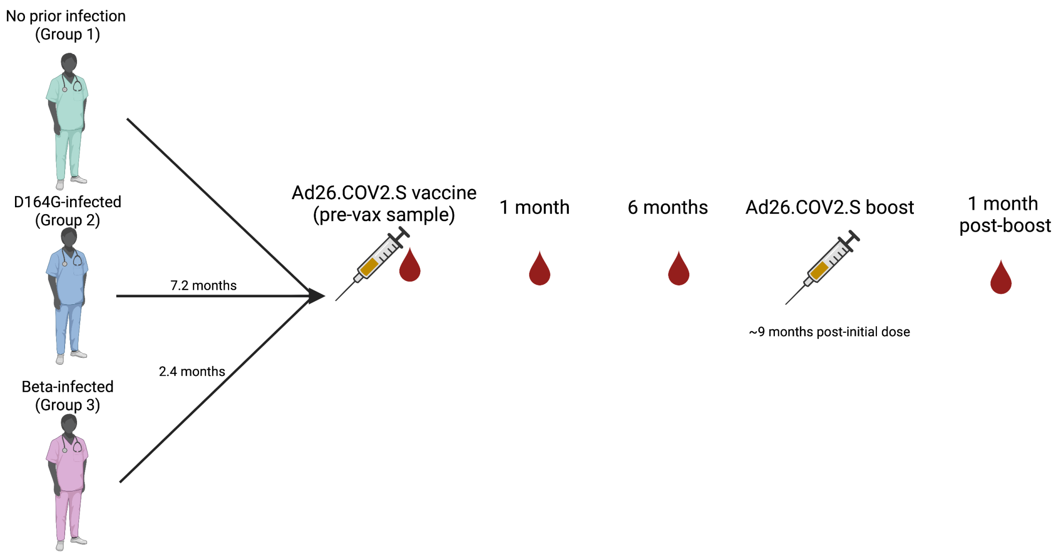


B

| | Longitudinal cohort | | |
|---|---|---|---|
| | No prior infection | D614G infected | Beta infected |
| N | 13 | 14 | 16 |
| Age (median, IQR) | 54 [35-58] | 35 [29-37] | 38 [33-48] |
| Gender (% female) | 77% | 64% | 75% |
| Time after initial vaccine dose (months, IQR) | 5.9 [5-6.3] | 5.3 [5.1-6.2] | 5.4 [5.1-6.1] |

**Supplementary Figure 1**

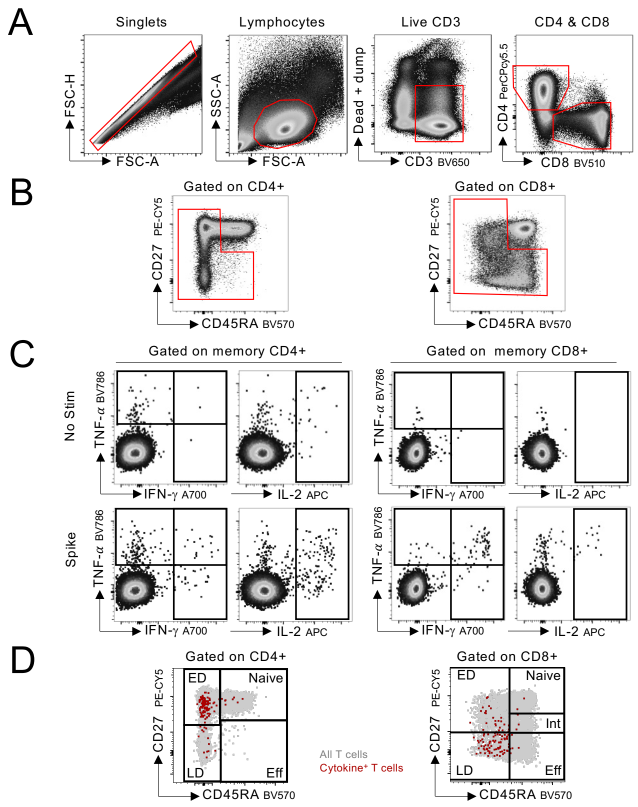


**Supplementary Figure 2**

### Binding

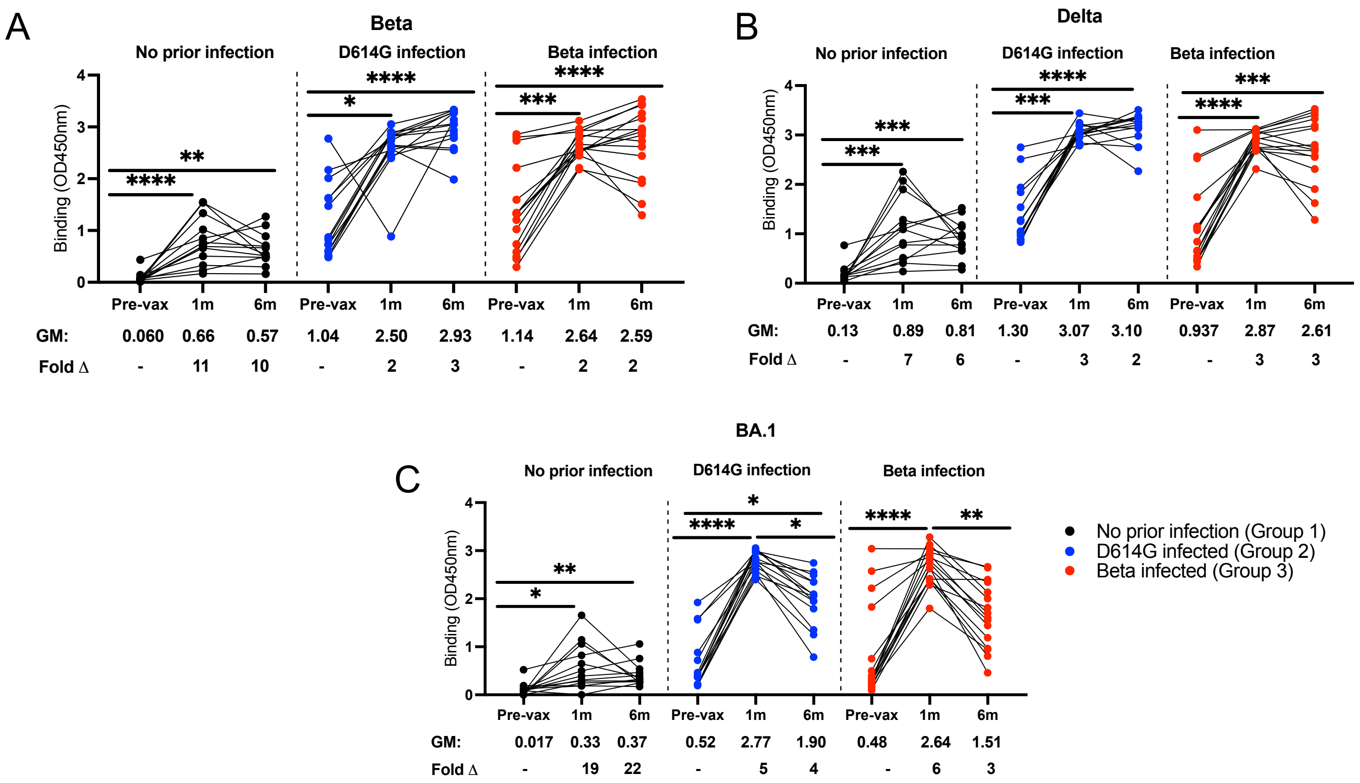


**Supplementary Figure 3**

### Antibody dependent cellular cytotoxicity (ADCC)

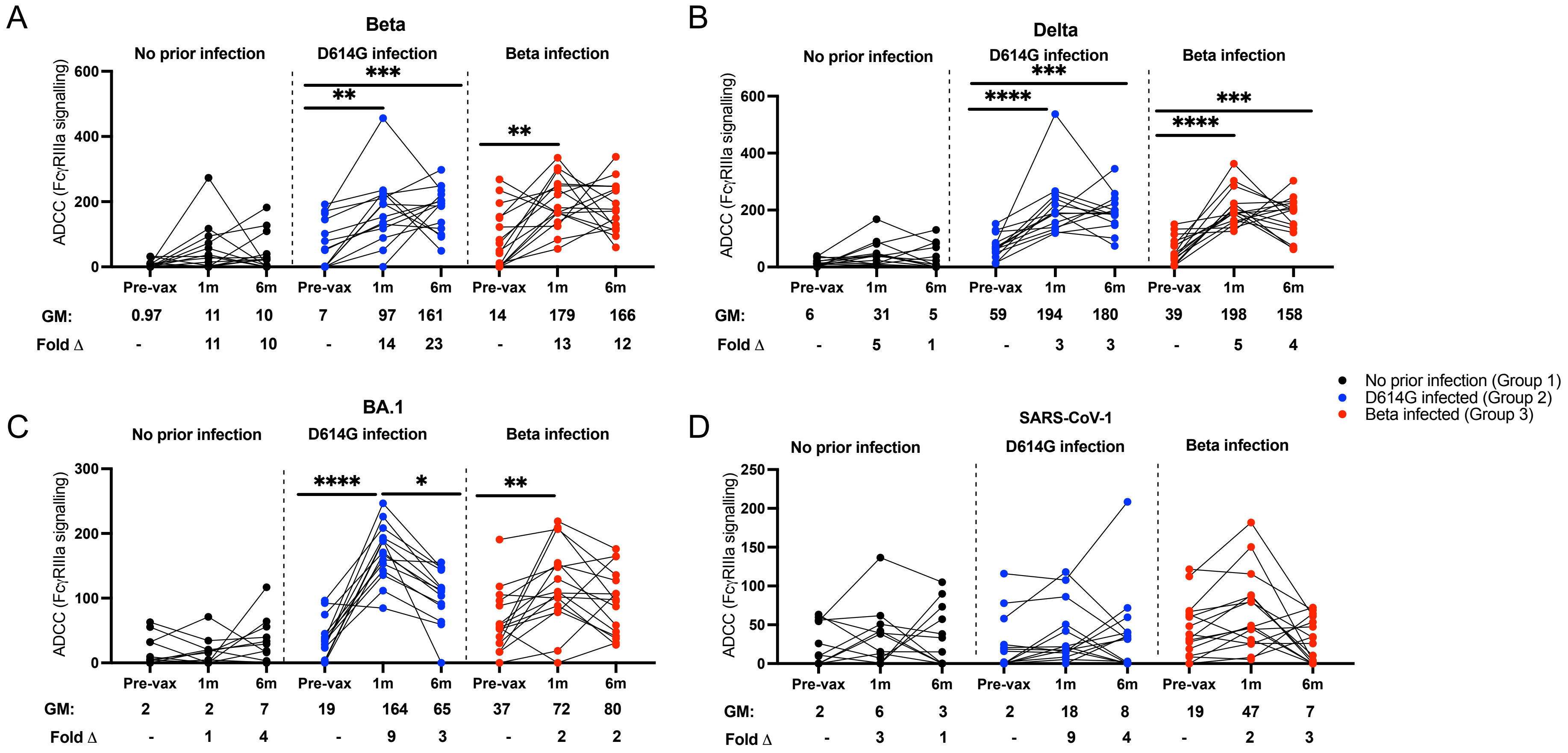


**Supplementary Figure 4**

### Neutralization

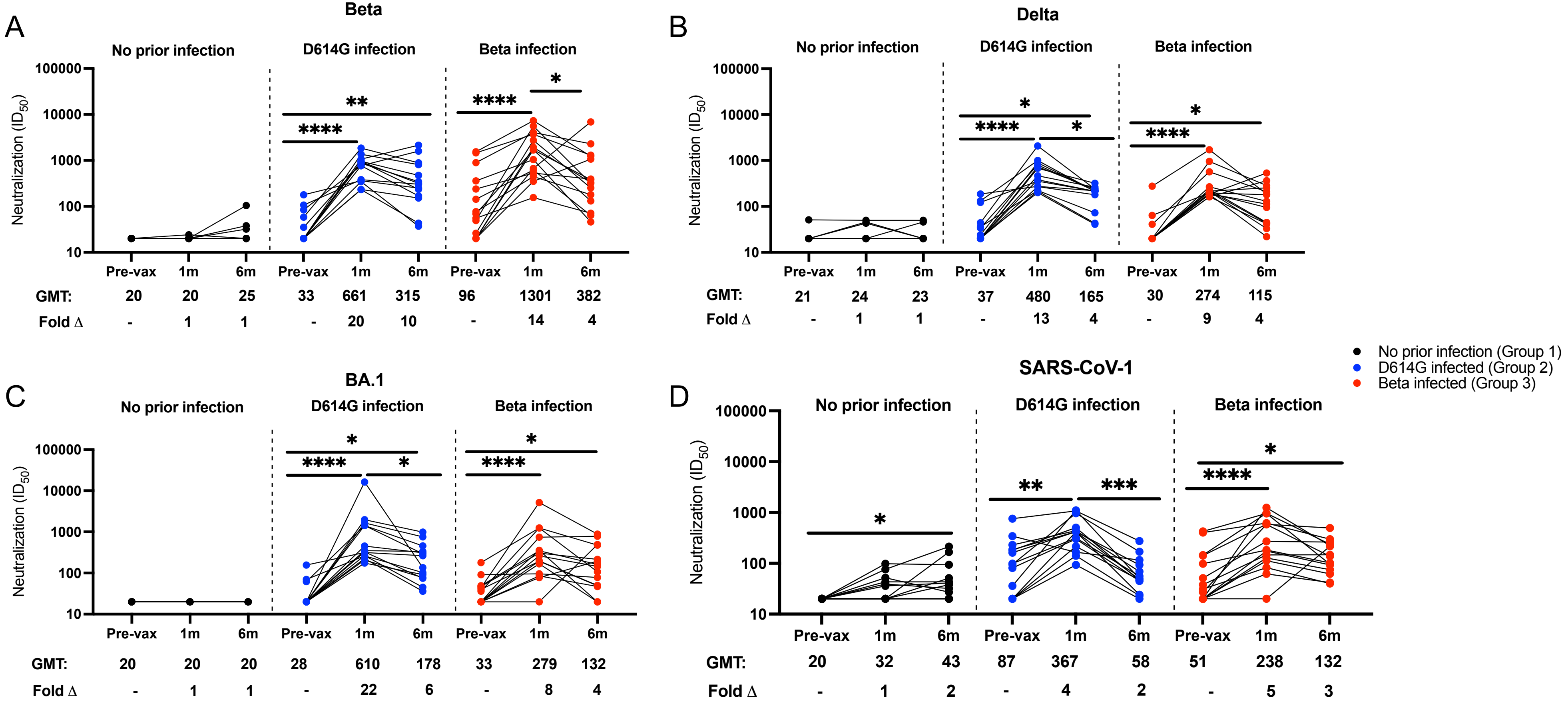


**Supplementary Figure 5**
